# Estimation of undetected COVID-19 infections in India

**DOI:** 10.1101/2020.04.20.20072892

**Authors:** Siuli Mukhopadhyay, Debraj Chakraborty

**Affiliations:** Department of Mathematics, Indian Institute of Technology Bombay, India; Department of Electrical Engineering, Indian Institute of Technology Bombay, India

**Keywords:** Discrete time, infection model, infection fatality rate, lockdown

## Abstract

**Background and Objectives:** While the number of detected COVID-19 infections are widely available, an understanding of the extent of undetected COVID-19 cases is urgently needed for an effective tackling of the pandemic and as a guide to lifting the lockdown. The aim of this work is to estimate and predict the true number of COVID-19 (detected and undetected) infections in India for short to medium forecast horizons. In particular, using publicly available COVID-19 infection data up to 28th April 2020, we forecast the true number of infections in India till the end of lockdown (3rd May) and five days beyond (8th May).

**Methods:** The high death rate observed in most COVID-19 hit countries is suspected to be a function of the undetected infections existing in the population. An estimate of the age weighted infection fatality rate (IFR) of the disease of 0.41%, specifically calculated by taking into account the age structure of Indian population, is already available in the literature. In addition, the recorded case fatality rate (CFR= 1%) of Kerala, the first state in India to successfully flatten the curve by consistently reporting single digit new infections from 12-20 April, is used as a second estimate of the IFR. These estimates are used to formulate a relationship between deaths recorded and the true number of infections and recoveries. The estimated undetected and detected cases time series based on these two IFR estimates are then used to fit a discrete time multivariate infection model to predict the total infections at the end of the formal lockdown period.

**Results:** Over three consecutive fortnight periods during the lockdown, it was noted that the rise in detected infections has decreased by 8.2 times. For an IFR of 0.41%, the rise in undetected infections decreased 2.5 times, while for the higher IFR value of 1%, undetected cases decreased by 2.4 times. The predicted number of total infections in India on 3rd May for both IFRs varied from 2.8 - 6.8 lakhs.

**Interpretation and Conclusions:** The behaviour of the undetected cases over time effectively illustrates the effects of lockdown and increased testing. From our estimates, it is found that the lockdown has brought down the undetected to detected cases ratio, and has consequently dampened the increase in the number of total cases. However, even though the rate of rise in total infections has fallen, the lifting of the lockdown should be done keeping in mind that 2.3 to 6.4 lakhs undetected cases will already exist in the population by 3rd May.

---

In this article, we propose a discrete time multivariate infection model for predicting the total true number of COVID-19 infections for a short to medium forecast horizon. Using publicly available COVID-19 data for India up to 28th April 2020, we predict the true number of infections in India during and up to the end of lockdown (3rd May 2020) period. For successful prediction of the total infections, we require an estimate of the extent of cases escaping detection. An estimate of the infection fatality rate (ratio of total deaths to total infections) for the Indian population has been calculated recently (Bommer and Vollmer (2020)). Assuming this rate to be constant, we determine estimates of the undetected infections for each day during the lockdown period. This time series data of undetected infections, along with recorded data for infected, recovered and deceased cases available from https://www.covid19india.org/, is used to fit a multivariate discrete time auto regressive (AR(1)) infection model. Using data up to 28th April, this model is used to predict both to-be-detected and to-be-undetected infections in India up to 3rd May and beyond.

The low detected infection numbers reported by a densely populated country like India is a highly debated subject with no known clear explanation. Many sources are attributing the low infection rates to the low number of tests being conducted for a country with 1.38 billion population. Bommer and Vollmer (2020) assessed the Indian COVID-19 data and suggested that India is detecting only 1.45% of the total number of infections. While, Srinivas and James (2020) concludes a 3.6% detection rate with wide variation among the states. Their assessment of low detection rate is based on the fact that the Indian case fatality rate (CFR = total number of deaths divided by total detected infections ≈ 10%) is poorly estimating the true infection fatality rate, due to a large number of undetected infections in India. Using a recent study by Verity et al. (2020) based on age stratified fatality data from mainland China and international Wuhan residents returning on repatriation flights, Bommer and Vollmer (2020) calculate an infection fatality rate (IFR) of 0.41% specifically for India. The IFR was calculated for India using population data from UN to correct for differences in age distributions in China and India. In our study we propose to use 0.41% as the first estimate (or lower estimate) of the IFR.

We obtain a second estimate of the IFR for India from cumulative death and infections data for the state of Kerala. Kerala has been exceptionally successful in reducing the number of new infections during the period between 12–20 April. Since the number of cases in Kerala at the beginning of the epidemic in India was quite high (e.g. maximum among all Indian states in March), this could only have been possible through successful tracking and isolation of almost every COVID-19 infection in the state. From this observation we argue, that Kerala had a very small number of undetected infections during that period. Now, the total deaths in Kerala up to 28th April (= 4) divided by the total confirmed cases up to 14th April (= 387) determines the CFR for Kerala to be 4/387 ≈ 1% on April 28. The reason for the fourteen day lag used in this calculation is explained in the next section. Due to our argument that the total reported infections in Kerala up to 14th April is approximately equal to the true number of total infections in the state up to that date, we assume the IFR to be approximately equal to this particular value of CFR. Consequently, this number (1%) is used as our second estimate (or upper estimate) of national IFR.

Weproposea2-equation discrete time AR(1)/state space model for the infection, death and recovered population dynamics. This is similar to the discrete time SIR model of Allen (1994) however with two major changes. Firstly, we ignore the *S* (susceptible population) equation. Since *S* is significantly larger as compared to current infection proliferation, we assume *S* is almost constant. Secondly, we let the coefficients of the model to vary linearly with time. We use these coefficient variations to model various forms of interventions such as lockdown, increased testing, etc.

## Materials and Methods

The prediction of total infections till the end of lockdown is broken into two steps. Firstly, we estimate the total number of undetected cases using the IFR for the period for which data on deaths are available. In the second step, this time series data of undetected infections, along with recorded data for infected, recovered and deceased are used in a multivariate discrete time auto regressive (AR(1)) infection model for predicting the total infections into the future.

*Step 1: Estimation of undetected cases based on IFR estimates up to the present time*: We describe an estimation procedure for the total number of undetected infections up to present time *t* (in our case 28th April). We denote,

*I_t_*: detected infections up to time *t*,
*D_t_*: deaths up to time *t*
*R_t_*: recoveries up to time *t*
*A_t_*: undetected infections up to time *t*.

Thus, the total number of infections recorded till time *t* is (*I_t_* + *A_t_*). Note, from our data set, we have observed values of *I_t_* and *D_t_*. However *A_t_* is unknown for all values of *t*.

Verity et al. (2020) reports that the average time from onset of symptoms in COVID19 to death to be approximately 18 days. As in Bommer and Vollmer (2020), we too allow a 4 day window from the start of symptoms for an infected individual to be recorded as a confirmed case. Thus, assuming that the total deaths till *t* results from the total number of infections recorded up to two weeks prior to *t*, we use the following relation to calculate *A_t_*_−14_:

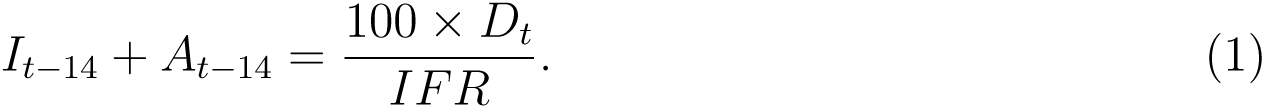

However, this formula cannot be used directly for the fourteen days preceding the present day, since we do not yet have the corresponding *D_t_* values. For these dates, we utilise the time variation of the ratio *A_t_/I_t_*, denoted by *K_t_* hereafter, to estimate *A_t_*. In the Results section we see that an exponential fit to *K_t_*, denoted by 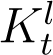, gives a good approximation of the variation in *K_t_*. Using the fitted 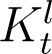 and *I_t_* from our data set, estimates of *A_t_* can be computed up to the present time.

*Step 2: Proposed Infection Model for Future Prediction of Undetected Cases*: The previous step provides us with estimates of *A_t_* up to present time. However, we would like to predict the number of cases in the near future (in general, for at least a 10 days horizon and in particular, up to the end of lockdown - 3rd May). To make this possible, we need to have a method for predicting *I_t_*, *D_t_* and *R_t_*. For reasons mentioned in the Introduction, for this short prediction horizon we prefer not to use a conventional SIR/SEIR type epidemiological model.

We model the relationship between the total number of infections (detected and undetected cases) and total deaths and recovered counts, based on the principle that the new infections, new deaths and new recoveries on day *t* + 1 is a (possibly time-varying) fraction *β_t_* of the infections active on the previous day *t*. In our notation, the new infections on day *t* + 1 is given by 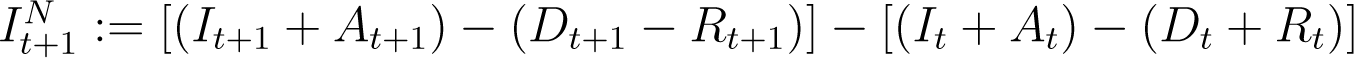. The new deaths and recoveries on day *t* + 1 is directly 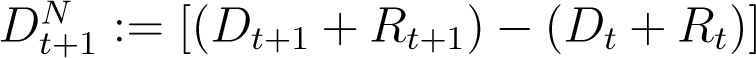, while the number of active infections on day *t* is 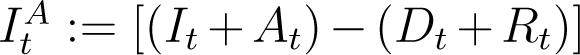. Hence from our hypothesis above,

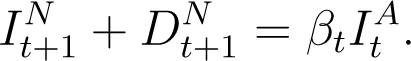

This equation simplifies to

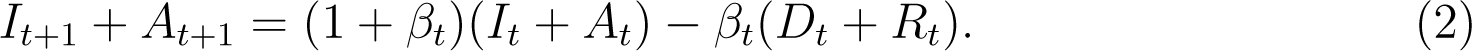

Note that the state variable *I_t_* + *A_t_*, is the cumulative count of total infections (active + dead + recovered) as opposed to cumulative active infections, which is frequently used in discrete time epidemiological models (Allen 1994). Due to our use of the IFR to estimate *A_t_* we find this version more convenient.

Similarly it is assumed that the new deaths and recoveries on day *t*+1 is a time varying fraction *γ_t_* of the number of active infections on day *t*:

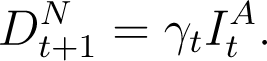

This equation simplifies to

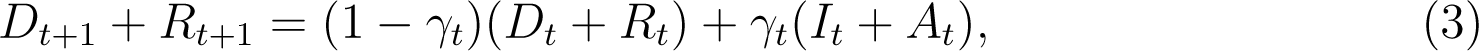

In principle, (2) and (3) can be used to simulate and predict the evolution of infections from any time onwards. However, just as the total number of infections is a sum of detected and undetected infections (*I_t_* + *A_t_*), the total recoveries at any time *t* is a sum of detected and undetected recoveries, which we denote by 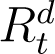 and 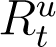 respectively. Hence,

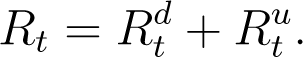

In (2) and (3) above, while 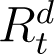 is recorded and known, 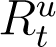 is unknown. Verity et al. (2020) states that the average time from onset of symptoms to recovery for individuals suffering from severe infections of COVID-19 to be 24 days, while from the WHO (2020) report the average recovery time for mild cases is 14 days. We assume individuals whose recoveries are enumerated on national databases of COVID-19 and are counted in 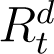, are likely to have had a severe infection, and consequently would have recovered in 24 days. Conversely, whose recoveries go undetected (as a part of 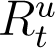) are likely to have suffered a milder form of the disease and would have recovered in 14 days from the start of symptoms. Allowing a four day window from the start of symptoms for each individual to be recorded as a confirmed/recovery case, and using using (1), we get,

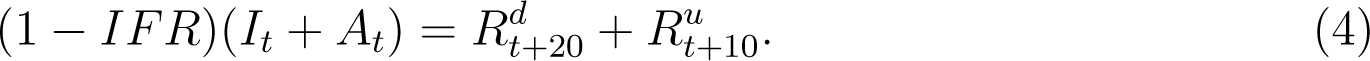

The above equation follows from the argument that if an infected individual did not die, he/she must have recovered in 10 or 20 days based on the severity of their infections. Using (4), we can calculate 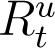 for all times except the first ten (*I_t_*_−10_, *A_t_*_−10_ not available) and the last ten samples (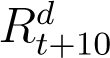 unavailable). While the first ten samples are unimportant, we need the last ten values of 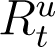 for further computation. This issue is resolved by studying the trend in the ratio 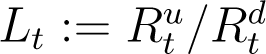 and fitting an exponential curve (denoted by 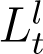) to the available data points. Using 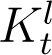, 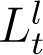 and available data, the model parameters are estimated as described next.

The assumption of time varying model coefficients *β_t_* and *γ_t_* is necessary for a good model fit to the data in the current scenario of rapidly evolving interventions such as lockdown/increased testing etc. Using the relations *A_t_* = *K_t_I_t_* and 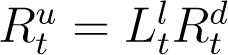, we can express *β_t_* and *γ_t_* as,

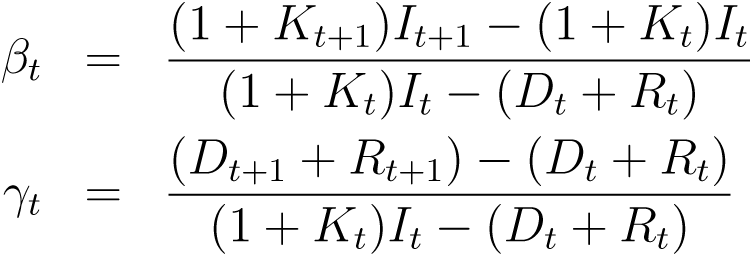

The values of *β_t_* and *γ_t_* for each *t* over a moving window of 10 days preceding the present day are plotted and linear curves are adaptively fitted to both *β_t_* and *γ_t_* over this moving window. Using these fitted values, denoted by 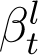 and 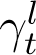, we find the predicted total number of infections (*I_t_* +*A_t_*) and total deaths and recoveries (*D_t_* +*R_t_*) by iterating (2) and (3) starting from 10 days in the past, up to the desired forecast horizon into the future.

## Results

### Results from Step 1

As explained in the Introduction, we use two IFR estimates. The first estimate is taken as 0.41% from Bommer and Vollmer (2020). The CFR of Kerala on 28 April (= 1%), is taken as the second estimate of the national IFR.

Based on these two IFRs, *A_t_* values for *t* (4th March - 14th April) are obtained directly from (1) and using *D_t_* recorded up to 28th April. Ratiosof *A_t_/I_t_*(= *K_t_*) over time *t* (4th March - 14th April) for both IFRs are shown in Figure 1. However, (1) cannot be used directly for 15th to 28th April since we do not yet have the corresponding *D_t_*+14 values. Hence, we utilise the variation in the ratios *A_t_/I_t_* with time to estimate *A_t_* for 15 - 28th April.

**Figure 1:**
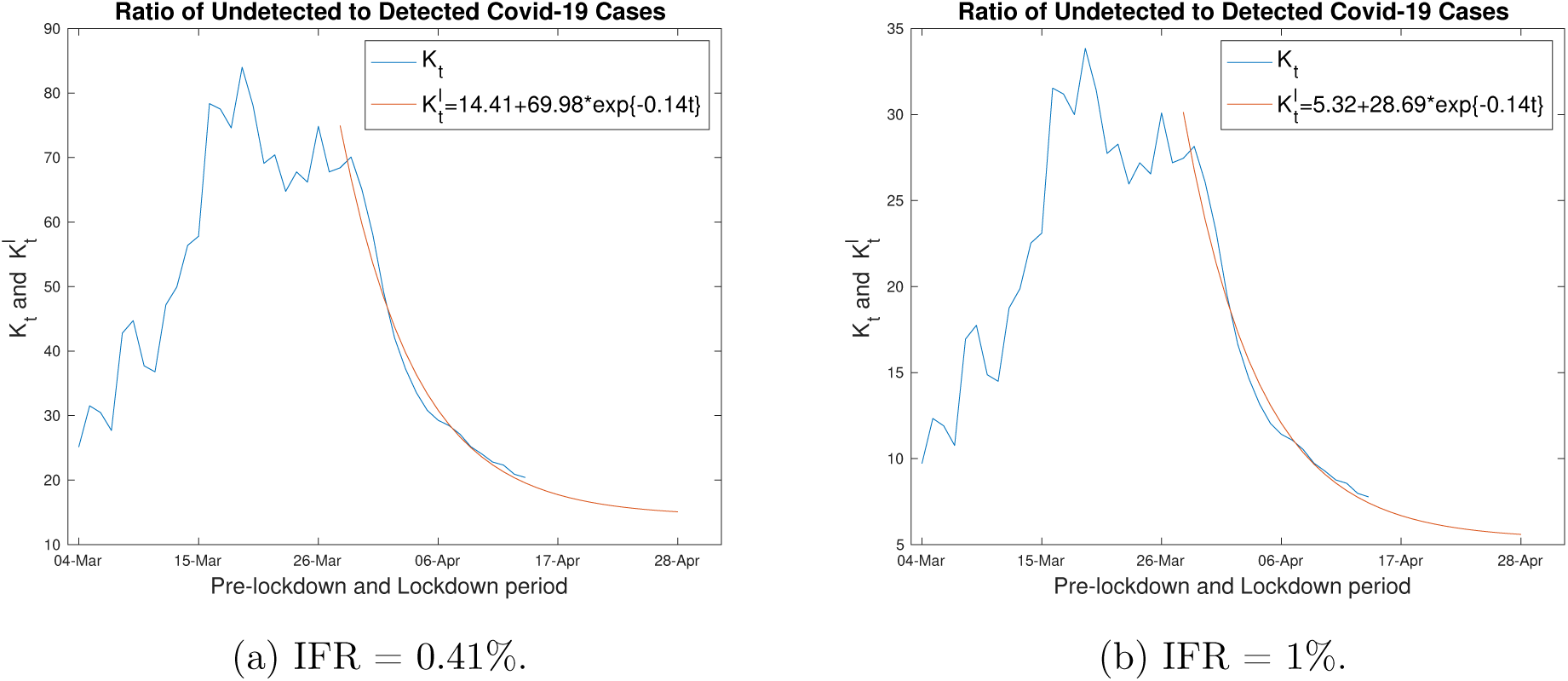
Estimated Ratio *A_t_/I_t_* and exponential curve fit

From Figure 1, it is quite clear that the nature of the plot differs before (increasing with time) and during the lockdown period (decreasing with time after 29th March). Since, we are interested in understanding the effect of the lockdown period, we fit an exponential curve 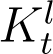 to *K_t_* from 29th March to 14th April. The fitted 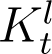’s (shown by the red lines in Figure 1) indicate exponentially decreasing ratios of undetected to detected infections in the lockdown period. The data appears to indicate that though the lockdown is effective in decreasing the infections it may not be able to completely eradicate the number of undetected infections. Hence we choose an exponential curve with an intercept to fit the *K_t_* values. As can be seen from Figure 1, the fitted 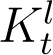 is a good approximation for *K_t_* after March 29th, and even after 5 April when *K_t_* starts to flatten out. This observation seems to indicate that the ratio of undetected to detected cases might hold constant or decrease only marginally up to the end of lockdown.

Using the exponentially fitted 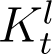 and the recorded *I_t_* from our data, we estimate *A_t_* for 15 - 28th April. These estimated undetected cases and the observed detected cases from 28th March till 28th April are plotted in Figure 2 for both IFRs.

**Figure 2:**
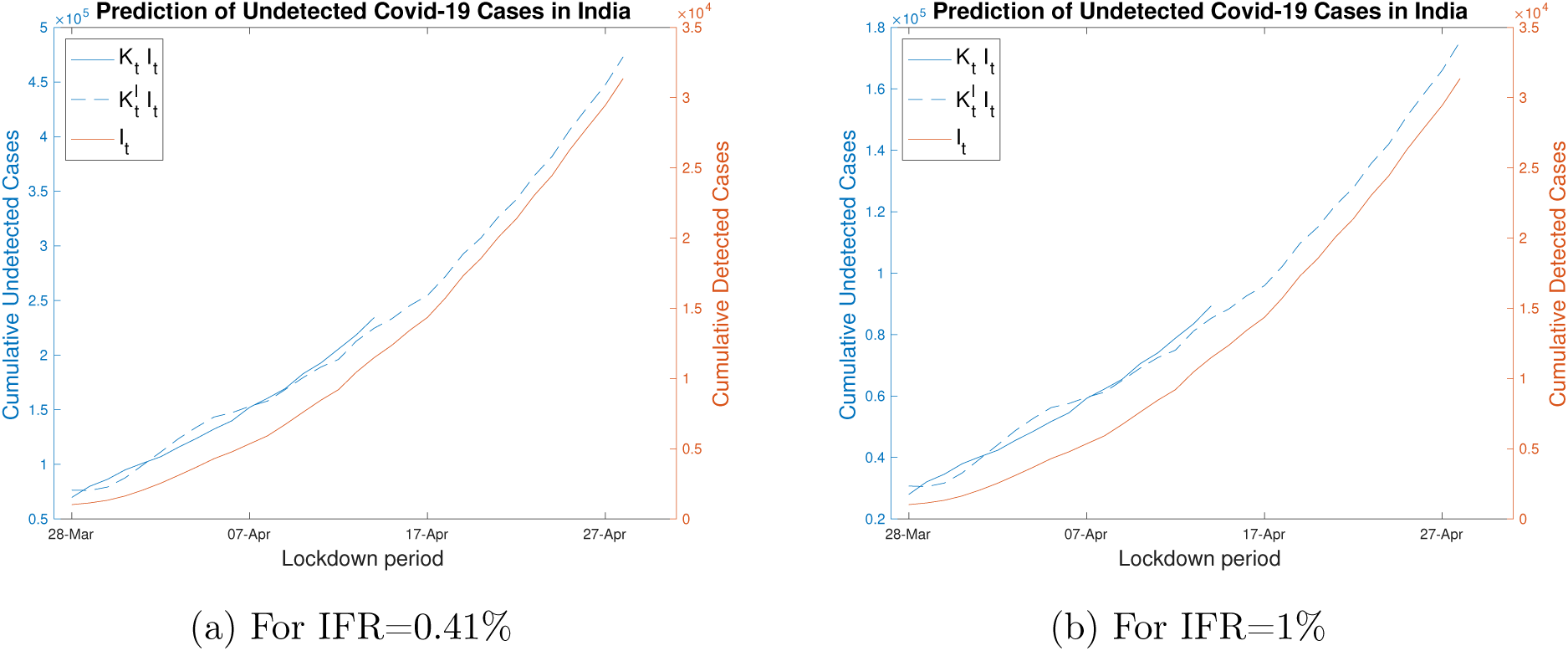
Detected cases *I_t_* (from data set) and undetected cases *A_t_*, estimated using *K_t_* and 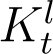. Note the difference in scales for *I_t_* and *A_t_*.

### Results for Step 2

We find the values of 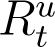 using (4). The ratios 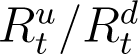 are plotted in Figure 3 for both IFRs. From the figure, we note that this ratio exhibit a similar behaviour (with a lag of 10 days) as the ratio of *A_t_/I_t_*. This is expected since the relatively high values of *A_t_* and 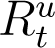, compared to *I_t_* and 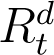 respectively, dominate these ratios, and through (4), 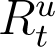 lags *A_t_* by 10 days. Recall that 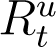 cannot be computed directly from (4) for the last 10 days preceding present time. As explained above, an exponential curve (with an intercept) is fitted to the available data points for 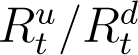 starting from 7th April (shown in red in Figure 3). This date is chosen since the data for 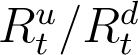 shows an exponential decay (similar to *K_t_* for 29th March) from that day onwards.

**Figure 3:**
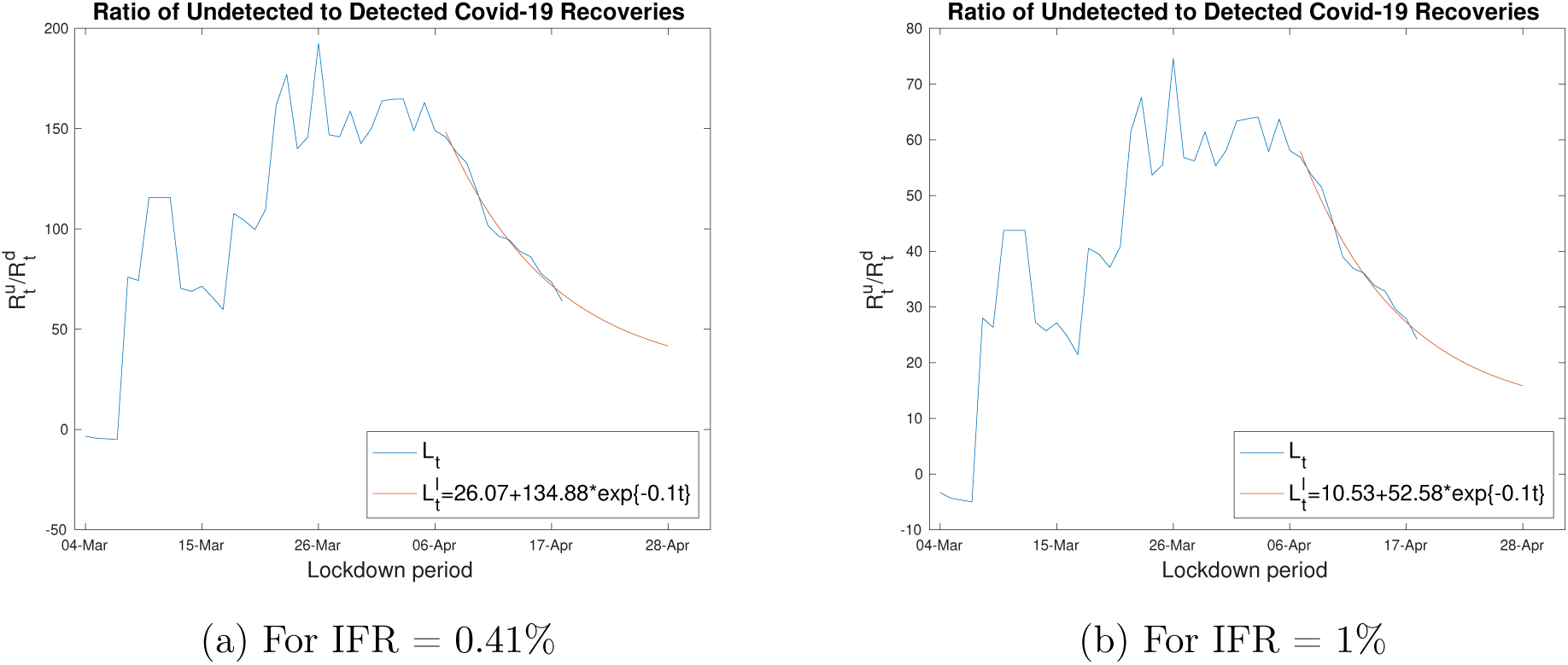
Estimated Ratio 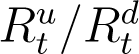 and exponential curve fit

The values of *β_t_* and *γ_t_* for 10 days preceding the present time (18 - 28 April) are plotted in Figures 4a and 4b respectively, for IFR of 0.41%. We adaptively estimate *β_t_* and *γ_t_* for both IFRs, by using the most recent disease data with a moving window of 10 preceding days. This is done to capture the most recent patterns in the disease data while estimating *β_t_* and *γ_t_*. The window size of 10 days was arrived at by trial and error. A larger window size cannot capture quickly changing trends in *β_t_* or *γ_t_*, while a very small window gives noisy estimates of both. A 10 day window is also justifiable since it is the minimum number of days required for effects of policy changes to be recorded in any data series (death, recovered or infection counts). The standard errors and confidence bounds for estimation of *β_t_* and *γ_t_* are reported in Table 1. Using these confidence bounds we can find upper and lower estimates for our predicted infection counts. To maintain simplicity, the bounds for the infected counts are however not shown here.

**Figure 4:**
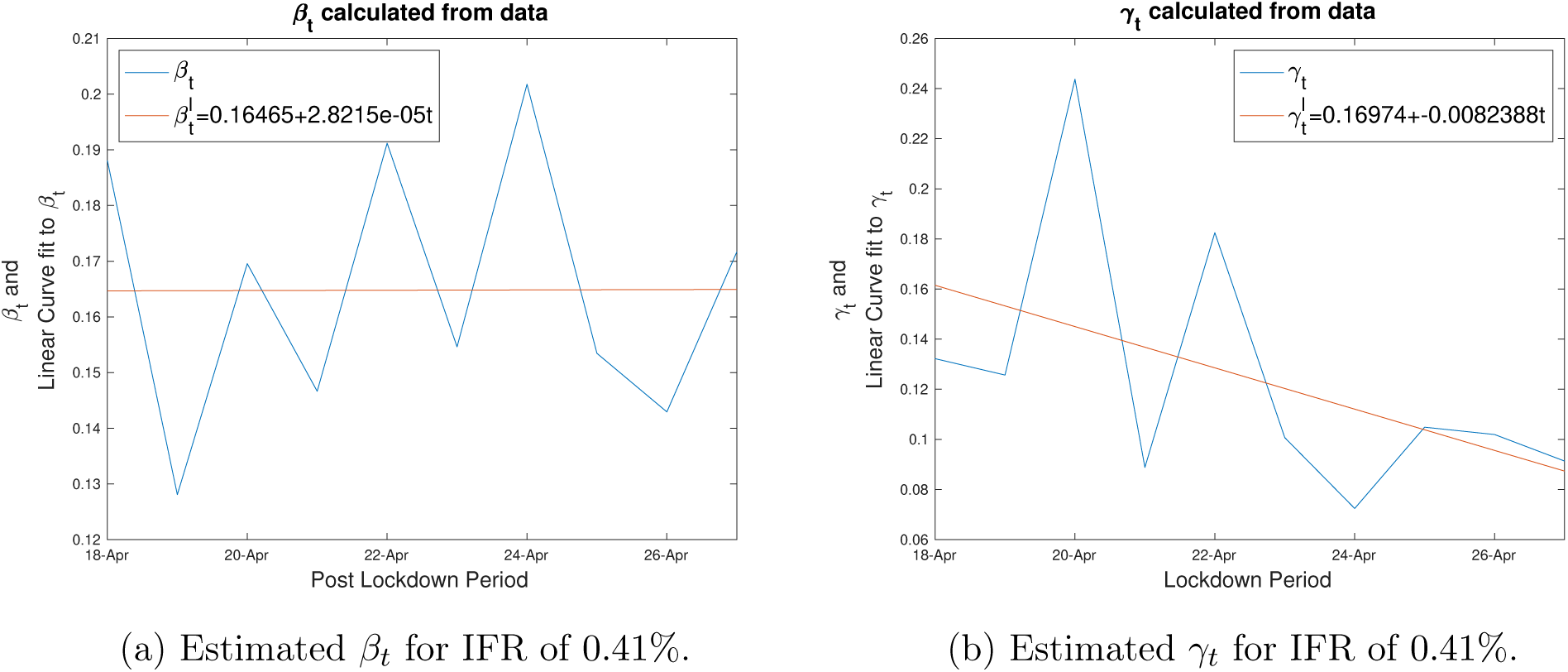
Model Parameter estimates over the preceding 10 day window

**Table 1:** Estimates and confidence bounds for 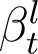 and 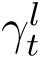 using an IFR of 0.41%. RMSE represents root mean squared error.

| Fitted Model | Coefficients | Estimates | Confidence bounds | RMSE |
| --- | --- | --- | --- | --- |
| $\beta_t^l$ | $b_1$ | $2.82 \times 10^{-5}$ | $(-0.0063, 0.0064)$ | 0.025 |
| | $b_2$ | 0.1646 | $(0.1250, 0.2043)$ | |
| $\gamma_t^l$ | $b_1$ | -0.0082 | $(-0.0205, 0.0040)$ | 0.0482 |
| | $b_2$ | 0.1697 | $(0.0938, 0.2457)$ | |

Using 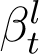 and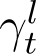, the proposed model (2) and (3) is initialized with the measured values of *I_t_, A_t_, D_t_* and *R_t_* on 18th April and iterated up to 8th May to simulate each variable up to this time for both values of IFR. The predicted values are plotted in Figures 5 and 6 for IFRs 0.41% and 1%, respectively, along with observed data up to 28th April. The summary statistics of the model fit and predictions are shown in Tables 2 and 3, for IFRs of 0.41% and 1% respectively. From the model fitting and prediction, the important findings can be summarised as:

- From Tables 2, using an IFR of 0.41%, we note that the total number of predicted infections (detected plus undetected) at the end of the lockdown period (3rd May) is 6.8 lakhs.
- From Table 3, using the higher IFR value of 1%, we note that the total number of predicted infections at the end of the lockdown period (3rd May) are 2.8 lakhs.
- In the first 14 day period (22nd March - 5th April) of the lockdown, there is a 10.7 times increase in the detected infections. The increase reduces to 2.5 times in the last fortnight period (19th April - 3rd May).
- For both IFR values, there was at least 4.5 times increase in undetected infections over the first fortnight into lockdown. However, in the last two weeks of lockdown the increase reduced to 2.1 times. These numbers indicate the rise in undetected infections have decreased due to the lockdown and testing intervention effect.
- The percentage of detections increased from 1% to 6% during lockdown for an IFR of 0.41%. Elevating the IFR to 1%, the rate of detection grew from 3% to 15%.
- It is noted from Figures 5 and 6 that the linear parameter varying model matches the data satisfactorily. Significantly, the proposed model can reproduce the sub-exponential growth of infection numbers observed over the lockdown period in India.

**Figure 5:**
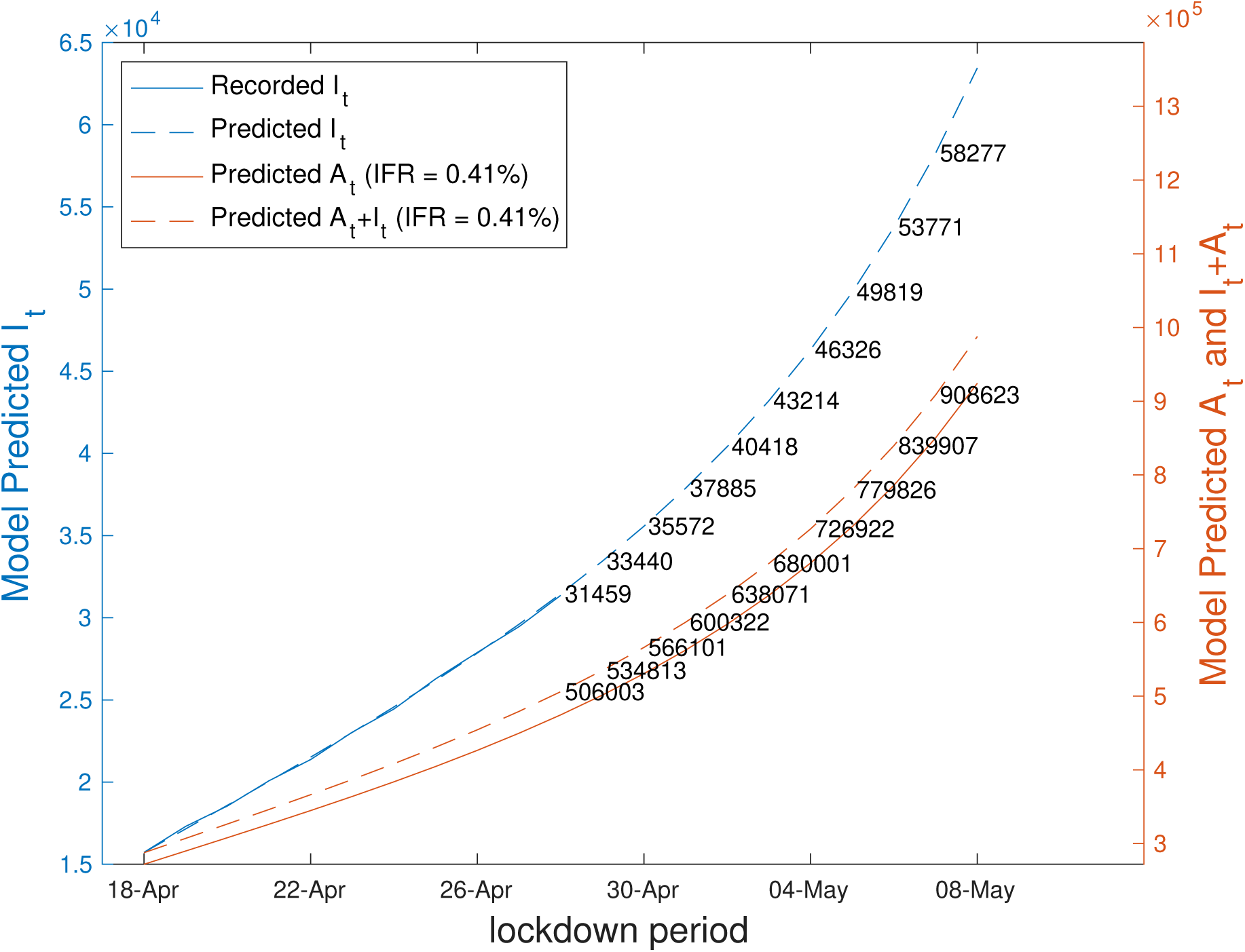
Predicted values of *A_t_*, *I_t_*, *I_t_* + *A_t_* and observed values of *I_t_* for IFR 0.41%. The predictions are obtained from (2).

**Figure 6:**
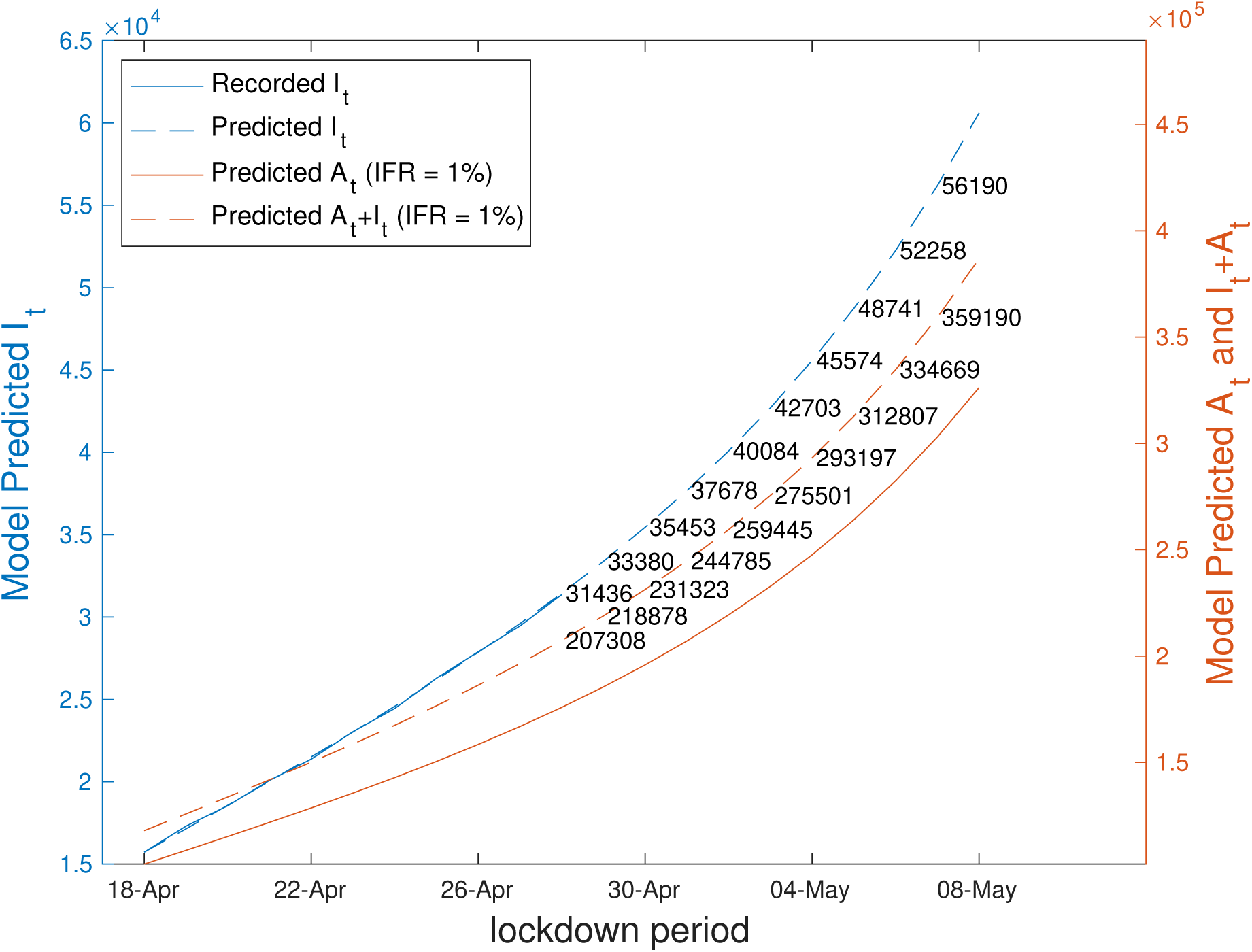
Predicted values of *A_t_*, *I_t_*, *I_t_* + *A_t_* and observed values of *I_t_* for IFR 1%. The predictions are obtained from (2).

**Table 2:** Predicted Values of *A_t_*, *I_t_* and *A_t_* + *I_t_* and ratio of detections using an IFR of 0.41%. The times increase are given in parenthesis.

| $t$ | $I_t$ | $A_t$ | $A_t + I_t$ | $I_t/I_t + A_t$ |
| --- | --- | --- | --- | --- |
| 22-Mar | 403 | 28377 | 28780 | 0.014 |
| 05-Apr | 4293 (10.7) | 132292 (4.7) | 136585 | 0.0314 (4.7) |
| 19-Apr | 17305 (4) | 292492 (2.2) | 309797 | 0.0559 (2.3) |
| 03-May | 43214 (2.5) | 636787 (2.2) | 680001 | 0.0635 (2.2) |

**Table 3:** Predicted Values of *A_t_*, *I_t_* and *A_t_* + *I_t_* and ratio of detections using an IFR of 1%. The times increase are given in parenthesis.

| $t$ | $I_t$ | $A_t$ | $A_t + I_t$ | $I_t/I_t + A_t$ |
| --- | --- | --- | --- | --- |
| 22-Mar | 403 | 11397 | 11800 | 0.0342 |
| 05-Apr | 4293 (10.7) | 51707 (4.5) | 56000 | 0.0767 (4.7) |
| 19-Apr | 17305 (4) | 109711 (2.1) | 127016 | 0.1362 (2.3) |
| 03-May | 42703 (2.5) | 232798 (2.1) | 275501 | 0.155 (2.2) |

## Discussion

The focus of our work is on predicting the total number of infections in India due to COVID-19 over a short forecast horizon and also studying the effect of lockdown and increased testing on these total infections.

The important observations from the first part of the Results section, are: From Figures 1 we see that in the pre-lockdown stage, the ratio 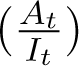 increased with time *t*. By 28th March the undetected cases were at least 60000 for an IFR of 0.41%. However, once the lockdown started, the ratio 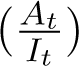 became almost constant over time. This slowing down effect on the rise in the undetected cases may be assumed to be the immediate effect of lockdown. From second week of the lockdown (29th March), the ratio 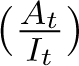 is noted to be decreasing with time. This leads to a further decrease in the number of undetected cases. This further decrease may be attributed to the additional effects of increased testing and active hotspot containment during the lockdown period. Thus, it seems from the data, that lockdown and increase in testing have lowered or slowed down the rate of rise in the number of undetected cases. Recently, however the decrease in this ratio seems to have saturated indicating that the current containment measures have reached their limits. From Figures 1a and 1b, we expect this ratio to hold constant or decrease only marginally until the end of the lockdown period. In addition, one should also note carefully the increasing *A_t_/I_t_* ratios prior to lockdown and consider the possible effect of increasing undetected cases, once the current intervention measures are relaxed. From the model fit and prediction in second step of the Results section, we note a fall in the increase in undetected cases and a simultaneous increase in detection of infections. At the end of the lockdown, we note that 42703 - 43214 infections will be detected while the undetected cases will vary between 2.3 - 6.4 lakhs, given the two IFR values. Thus, it seems lockdown and increased testing have been effective measures in reducing the rise in infections from COVID-19 in India. However, as a word of caution, we would like to add that though the rate of increase of undetected cases seems to have slowed down with the interventions, at the end of lockdown we will still have 2.8 - 6.8 lakhs total existing infections to combat.

We would also like to point out that the linear fit to the model coefficients (*β_t_* and *γ_t_*) are valid only in the short term, and the proposed infection model should not be used for long term predictions.

We understand that ignoring the susceptible population dynamics, limits the forecasting capabilities of the proposed model. However, we believe that this is a reasonable assumption in the short to medium term when the susceptible population remains very high and the recovered population is negligible. As reported above, we see a dampening in the exponentially increasing infection figures over differing time periods (during and up to the end of the formal lockdown period). We believe that this decrease in the infection rate is due to intervention measures and gradual buildup of awareness in the general population rather than development of herd immunity or recovery dynamics. As an extension of this work, we plan to use our model for predicting state-wise total infections.

## Data Availability

Publicly available at https://www.covid19india.org

## References

• Allen J S (1994). Some discrete-time SI, SIR, and SIS epidemic models. Mathematical Biosciences Volume 124, Issue 1, November 1994, Pages 83–105

• Bommer C and Vollmer S (2020) Average detection rate of SARS-CoV-2 infections is estimated around six percent. www.uni-goettingen.de/en/606540.html

• Goli S and James K S (2020) How much India detecting SARS-CoV-2 Infections? A model-basedestimation. medRxivpre print doi: https://doi.org/10.1101/2020.04.09.20059014.

• Verity et al. Estimates of the severity of coronavirus disease 2019: a model-based analysis. The Lancet Infectious Diseases 2020. DOI: 10.1016/S1473-3099(20)30243-7

• WHO 2020. Report of the WHO-China Joint Mission on Coronavirus Disease 2019 (COVID-19). https://www.who.int/docs/default-source/coronaviruse/who-china-jointmission-on-covid-19-final-report.pdf

